# Multi-Task Learning and Soft-Label Supervision for Psychosocial Burden Profiling in Cancer Peer-Support Text

**DOI:** 10.64898/2026.04.03.26350034

**Authors:** Zhongyan Wang, Yuchen Cao, Xiaorui Shen, Zhanyi Ding, Yunchong Liu, Yeyubei Zhang

**Affiliations:** Center for Data Science, New York University, New York, NY, USA; Khoury College of Computer Science, Northeastern University, Boston, MA, USA; School of Engineering and Applied Science, University of Pennsylvania, Philadelphia, PA, USA

**Keywords:** natural language processing, neoplasms, psychosocial support systems, machine learning, patient-generated health data

## Abstract

**Objective:** Online cancer peer-support text contains signals of psychosocial burden beyond emotional tone, including treatment burden, financial strain, uncertainty, and unmet support needs. We evaluated 2 modeling extensions: multi-task learning (MTL) for joint prediction of health economics and outcomes research (HEOR) burden dimensions, and soft-label supervision using large language model (LLM)-derived probability distributions.

**Materials and Methods:** We analyzed 10,392 cancer peer-support posts. GPT-4o-mini generated proxy annotations for HEOR burden subscales, composite burden, high-need status, speaker role, cancer type, and emotion probabilities. Study 1 trained a shared ALBERT encoder under 4 MTL conditions: composite and subscale burden targets, each with and without auxiliary heads, using Kendall uncertainty weighting. Study 2 compared soft-label training on LLM emotion distributions with hard-label baselines under regular and token-augmented inputs, evaluating performance against both human labels and AI distributions.

**Results:** Composite-only MTL achieved *R*^2^=0.446 for burden regression and weighted F1=0.810 for high-need screening; subscale classification achieved mean weighted F1=0.646. Adding auxiliary role and cancer-type heads reduced regression performance (Δ*R*^2^ = −0.209). Soft-label training reduced weighted F1 by 0.16 versus hard-label baselines (0.68 vs. 0.86), and token augmentation did not improve performance under soft supervision.

**Discussion:** Composite-only MTL supported modeling of multidimensional burden-related signals from forum text, whereas auxiliary prediction heads appeared to compete with primary tasks. Soft-label training aligned poorly with human-labeled emotion categories, suggesting that uncalibrated LLM distributions may propagate bias rather than improve supervision.

**Conclusion:** Composite-only MTL was the strongest burden-modeling approach, and hard-label supervision remained preferable for emotion classification.

## 1 BACKGROUND AND SIGNIFICANCE

Cancer survivors and caregivers frequently use online peer-support communities to describe treatment experiences, emotional distress, financial strain, uncertainty, and unmet support needs in their own words. ^1–3^ These patient-generated health data (PGHD) offer a scalable source of information on psychosocial burden that is often difficult to capture through structured clinical instruments alone. Prior natural language processing (NLP) studies in cancer support forums have focused mainly on sentiment or emotion classification, ^4–6^ showing that transformer-based models can identify broad emotional tone with moderate to strong performance. However, emotional valence alone provides only a partial view of psychosocial burden. Posts expressing financial stress, treatment toxicity, uncertainty about prognosis, or lack of support may all appear similarly negative while implying different support needs and potentially different intervention pathways.

This limitation motivates a shift from single-dimension sentiment classification toward more granular burden modeling. Health economics and outcomes research (HEOR) frameworks distinguish among burden dimensions such as treatment burden, cost burden, perceived harm, life disruption, uncertainty and decisional conflict, support and coping resources, and perceived benefit. ^7^ Mapping forum language to these dimensions could improve the usefulness of automated monitoring systems by moving beyond general negativity toward more specific burden-related signals. Yet, to our knowledge, prior cancer NLP studies have not jointly modeled multiple psychosocial burden dimensions from peer-support text.

Multi-task learning (MTL) ^8^ offers a natural way to address this problem. A shared encoder can learn common linguistic representations while predicting multiple related targets, potentially improving efficiency and enabling richer profiling from a single model. In clinical NLP, MTL is attractive when outcomes are conceptually related but differ in form, such as continuous scores, binary screening flags, and categorical subscales. A practical challenge, however, is that heterogeneous tasks can differ substantially in scale and difficulty. Learned loss-balancing methods such as homoscedastic uncertainty weighting ^9^ provide one solution by adaptively scaling task contributions during training. Even with such balancing, auxiliary tasks do not necessarily help: easier tasks may dominate shared optimization rather than improve representations for the primary outcome.

A second methodological question concerns label provenance. Large language models (LLMs) are increasingly used to annotate health text at scale, ^10,11^ making it feasible to derive labels for constructs that would otherwise be expensive to annotate manually. LLMs can also produce class-probability distributions rather than only hard labels. In principle, these “soft labels” may preserve uncertainty that argmax labels discard and may therefore provide a richer supervision signal. ^12^ In practice, however, LLM annotations may exhibit systematic bias or miscalibration on subjective tasks, ^13,14^ raising the possibility that soft-label training could propagate annotator bias rather than improve downstream performance. This concern is especially relevant when the ultimate evaluation target is human judgment rather than agreement with the LLM itself.

Our companion studies on this dataset established relevant baselines. Wang et al. ^15^ evaluated four-class emotion classification with human labels using ALBERT, and Xu et al. ^16^ showed that LLM-derived labels shifted the class distribution toward negativity while token augmentation with LLM-extracted metadata modestly improved hard-label classification. These findings raise 2 linked questions. First, can LLM-derived annotations support multi-dimensional psychosocial burden modeling beyond emotion classification? Second, can LLM probability distributions be used directly as soft supervision targets, or does their distributional bias degrade performance relative to hard-label training?

We address these questions through 2 complementary studies using the same corpus, encoder family, and split protocol. Study 1 develops a multi-task framework for modeling LLM-defined HEOR burden dimensions from cancer peer-support text. We compare composite-level prediction with subscale-level prediction and test whether auxiliary speaker-role and cancer-type heads improve or interfere with the primary burden tasks. Study 2 evaluates soft-label supervision for emotion classification by comparing training on LLM-derived probability distributions against hard-label baselines under regular and token-augmented inputs. Because Study 1 depends on LLM-generated burden annotations as task targets, Study 2 serves as a targeted audit of whether richer LLM probability outputs can also be trusted as supervision signals.

This work makes 4 contributions. First, it presents a unified empirical evaluation of multi-task burden modeling and soft-label supervision in cancer peer-support text. Second, it shows that composite-only MTL can model multidimensional burden-related signals from forum posts, whereas adding auxiliary prediction heads may reduce primary-task performance. Third, it shows that, in this setting, LLM-derived soft-label supervision underperforms hard-label training, highlighting the importance of calibration and label-quality assessment before using LLM probability distributions as training targets. Fourth, it demonstrates that token augmentation, modestly effective under hard-label supervision, provides no benefit under soft-label training, establishing that augmentation strategies should be evaluated conditional on label quality rather than as architecture decisions independent of the supervision source.

## 2 MATERIALS AND METHODS

### 2.1 Dataset

We used the *Mental Health Insights: Vulnerable Cancer Survivors & Caregivers* corpus ^17^ (*N* = 10,392 posts; CC BY 4.0), a publicly available collection of posts from an online cancer peer-support forum spanning multiple cancer types. Two human raters assigned each post an ordinal emotional intensity label on a 4-point scale (− 2=very negative, −1=negative, 0=neutral, +1=positive). Class frequencies were 4,375 neutral (42.1%), 4,112 negative (39.6%), 1,155 very negative (11.1%), and 750 positive (7.2%). Median post length was 162 words (IQR, 93– 276).

Preprocessing included lowercasing, replacement of URLs, user mentions, and hashtags with generic placeholders, removal of non-alphanumeric characters, and whitespace normalization. Text was tokenized with the ALBERT tokenizer ^18^ (`albert-base-v2`) using a maximum sequence length of 200 tokens.

Data were split with a 2-stage stratified holdout procedure into training, validation, and test sets (60/20/20; random seed=42). Study 1 stratified on the binary high-need flag, whereas Study 2 stratified on the 3-class human emotion label. Thus, the two studies used identical split ratios and seed but not identical partitions. The same corpus and encoder family were used in companion studies by Wang et al. ^15^ and Xu et al. ^16^, supporting close cross-study comparison. This secondary analysis used publicly available de-identified data and did not require institutional review board approval (45 CFR 46.104(d)(4)).

### 2.2 LLM Annotation

GPT-4o-mini ^19^ generated all AI-derived labels under deterministic settings (temperature=0; structured JSON output; batches of 20 posts). Annotation was applied to the full corpus before data splitting and treated as a fixed preprocessing step. These outputs were treated as proxy annotations rather than validated clinical labels. In Pass 1, the LLM produced: (1) a 4-class emotion probability distribution over {very negative, negative, neutral, positive}; (2) speaker role (Patient, Caregiver, or Unclear); and (3) cancer type (brain, colon, liver, leukemia, lung, other, or unknown).

In Pass 2, the LLM generated 7 psychosocial burden subscales informed by health economics and outcomes research (HEOR): perceived benefit, perceived harm, cost burden, treatment burden, life disruption, uncertainty/decisional conflict, and support/coping resources. Each subscale was scored on a 4-point ordinal scale (0– 3). The LLM also generated a composite burden score (0–100) and a binary high-need flag. These HEOR labels were LLM-generated proxy annotations without human subscale validation.

This created a dual-annotation setting ^12^ in which human labels served as ground truth for emotion classification, whereas AI labels served as proxy burden targets for Study 1 and soft probability targets for Study 2.

### 2.3 Study 1: HEOR Multi-Task Learning

#### 2.3.1 Architecture

Study 1 evaluated whether a single transformer encoder could jointly predict multiple LLM-defined HEOR burden dimensions using multi-task learning (MTL). ^8^ We tested 2 design factors: HEOR granularity (composite vs. subscales) and auxiliary context heads (without vs. with speaker role and cancer type). Role and cancer type were modeled as auxiliary prediction heads rather than input tokens.

All models used ALBERT^18^ (`albert-base-v2`; 12M parameters) with task-specific heads attached to the [`CLS`] representation. Four MTL conditions were evaluated in a 2 *×* 2 design:

1. **Composite**: regression head for total burden score (MSE) and binary head for high-need flag (BCE).
2. **Composite+RC**: Composite plus speaker role (3-class CE) and cancer type (7-class CE).
3. **Subscales**: 7 separate 4-class heads, 1 for each HEOR subscale (class-weighted CE).
4. **Subscales+RC**: Subscales plus speaker role and cancer type.

Classification heads used dropout followed by linear projection; the regression head used a single linear unit.

#### 2.3.2 Loss balancing

Because MSE, BCE, and CE losses differ in scale, we used homoscedastic uncertainty weighting. ^9^ For task *t*, the joint loss was:

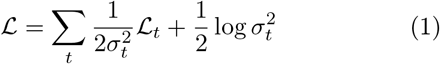

where 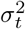 is a learned task-specific variance. Log-variance terms were initialized at 0 and optimized with learning rate 0.01 to allow rapid adjustment of relative task weights. In preliminary experiments, equal weighting yielded unstable performance in the Composite+RC condition, supporting the use of learned loss balancing in this setting.

#### 2.3.3 Hyperparameter search

For each Study 1 condition, we performed 10 random-search iterations over learning rate (1 × 10^−5^ to 3 × 10^−5^, log-uniform), epochs (6–9), and dropout (0–0.25). Model selection used the mean validation score across active heads. Early stopping used patience=3, and the best validation checkpoint was retained.

### 2.4 Study 2: Soft-Label Supervision

#### 2.4.1 Three-class collapse

Xu et al. ^16^ showed that 4-class LLM emotion distributions were diffuse (Jensen-Shannon divergence ^20^ = 0.1155 from human labels), particularly for the very negative class. We therefore collapsed very negative and negative into a single Negative class for Study 2. The AI soft-label vector was defined as *P* (Negative) = *P* (very negative) + *P* (negative), retaining *P* (Neutral) and *P* (Positive). Human labels were collapsed analogously: −2 and −1 to Negative, 0 to Neutral, and +1 to Positive. A supplementary 4-class soft-label analysis is also reported.

#### 2.4.2 Experimental conditions

We evaluated 2 input conditions under soft-label supervision: **Regular** (raw post text) and **Augmented** (post text prepended with `ROLE_<role> CANCER_<type>` tokens from Pass 1 annotations). Both conditions trained on the 3-class AI probability distributions using soft cross-entropy:

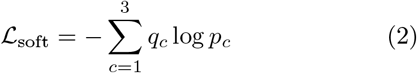

where *q*_*c*_ is the AI-derived probability for class *c* and *p*_*c*_ is the model prediction. Model selection used validation weighted F1 against human labels to preserve alignment with the ultimate evaluation target.

Test-time evaluation used 2 metric schemes: hard-metric evaluation against human 3-class labels (weighted F1, macro AUC, and per-class precision/recall/F1) and soft-metric evaluation against the AI probability distributions (soft cross-entropy and Brier score ^21^). Study 2 used the same ALBERT encoder and hyperparameter search protocol as Study 1. Hard-label baselines from Xu et al. ^16^ served as the primary comparison.

#### 2.4.3 Evaluation

All models were evaluated on the held-out test set (*n* = 2,079). In Study 1, composite conditions were evaluated with RMSE, *R*^2^, and MAE for burden regression and weighted F1 for the binary high-need flag. ^22^ Subscale conditions were evaluated with per-subscale weighted F1 and the mean across the 7 subscales. Auxiliary heads were evaluated with weighted F1.

In Study 2, hard-metric evaluation assessed alignment with human labels, whereas soft-metric evaluation assessed fidelity to the LLM probability landscape. ^23^ We estimated 95% confidence intervals using 2,000 stratified bootstrap resamples of the test set ^24^. Experiments were implemented in PyTorch ^25^ with Hugging Face Transformers ^26^.

## 3 RESULTS

Results are reported by study. Study 1 evaluated HEOR multi-task learning under 4 MTL conditions; Study 2 evaluated soft-label emotion classification under 2 input conditions (regular vs. augmented). All metrics were computed on the held-out test set (*n* = 2,079) with bootstrap 95% confidence intervals (2,000 resamples). Study 1 was evaluated against LLM-derived burden targets, whereas Study 2 was evaluated against human emotion labels unless otherwise noted.

### 3.1 Study 1: HEOR Multi-Task Learning

#### 3.1.1 Composite burden prediction

The Composite model achieved moderate regression performance for total burden score (RMSE=13.47, 95% CI: 12.95–14.00; *R*^2^=0.446, 95% CI: 0.403–0.487; MAE=10.15 on a 0–100 scale) and strong performance for high-need classification (weighted F1=0.810, 95% CI: 0.792–0.827), with high recall for high-need posts (0.935) and precision of 0.796 (Table 1). This recall-favoring operating point is appropriate for a screening application in which missing a high-need post carries greater cost than over-flagging for moderator review.

**Table 1.** Composite HEOR Regression and High-Need Classification.

| Metric | Composite | Comp.+RC | $\Delta$ |
| --- | --- | --- | --- |
| RMSE | 13.47 [12.95, 14.00] | 15.81 [15.29, 16.33] | +2.33 |
| $R^2$ | 0.446 [0.403, 0.487] | 0.237 [0.181, 0.292] | -0.209 |
| MAE | 10.15 | 12.40 | +2.25 |
| High-Need wF1 | 0.810 [0.792, 0.827] | 0.794 [0.775, 0.812] | -0.016 |
| Role wF1 | — | 0.913 [0.900, 0.924] | — |
| Cancer wF1 | — | 0.688 [0.667, 0.711] | — |

Deltas indicate the effect of adding auxiliary role and cancer-type heads. CI = bootstrap 95% confidence interval. wF1 = weighted F1.

Adding speaker role and cancer type as auxiliary prediction heads (Composite+RC) reduced performance on both primary tasks. *R*^2^ decreased from 0.446 to 0.237 (Δ = −0.209), and weighted F1 for high-need classification decreased from 0.810 to 0.794, despite strong role prediction (F1=0.913) and moderate cancer-type prediction (F1=0.688). Thus, the auxiliary heads were learnable, but their inclusion coincided with worse HEOR performance.

#### 3.1.2 Subscale burden prediction

Among the 7 HEOR subscales, cost burden had the highest weighted F1 (0.852, 95% CI: 0.836–0.867), followed by support/coping (0.692), treatment burden (0.627), benefit (0.624), life disruption (0.611), uncertainty (0.584), and harm (0.531, 95% CI: 0.511–0.551). Mean weighted F1 across subscales was 0.646.

Adding auxiliary heads (Subscales+RC) reduced mean subscale F1 to 0.624, with 6 of 7 subscales showing lower performance. The largest declines occurred for uncertainty (−0.060) and harm (−0.047); life disruption showed a small improvement (+0.021). In this condition, role prediction remained strong (F1=0.910), whereas cancer-type performance declined to F1=0.563.

#### 3.1.3 Learned task weights

The learned task weights (Table 2) provide one explanation for the auxiliary-task pattern. In Subscales+RC, role prediction received 3.97 of 9.0 total normalized weight units (44%), whereas cancer type received 0.33. A similar pattern appeared in Composite+RC, where role received 1.40 of 4.0 units (35%). Among HEOR tasks, cost burden and support/coping received the largest weights, whereas harm received the smallest.

**Table 2.**
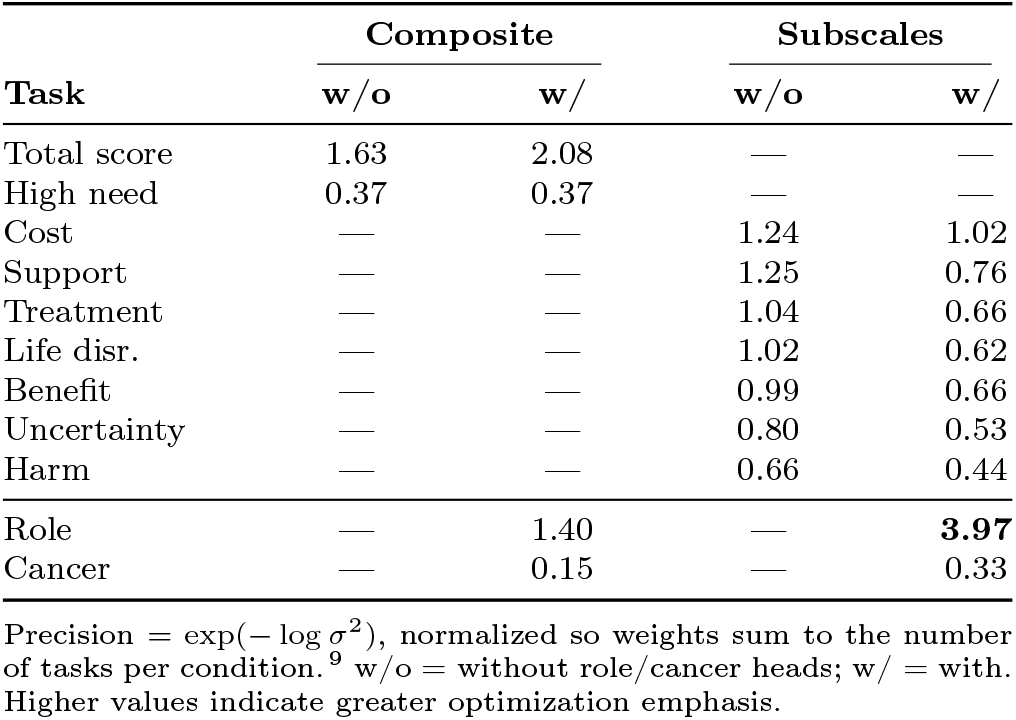
Kendall-Learned Task Weights (Normalized)

### 3.2 Study 2: Soft-Label Supervision

#### 3.2.1 Soft-label entropy

After collapsing the 4-class LLM emotion output into 3 classes, the resulting distributions showed lower but still substantial entropy. ^27^ Mean probability on the argmax class was 0.725 for Negative, 0.607 for Neutral, and 0.566 for Positive. Corresponding entropies were 0.710, 0.802, and 0.894 (maximum for 3 classes, ln 3 = 1.099). The Positive class remained highly diffuse, with entropy near 81% of the theoretical maximum.

#### 3.2.2 Classification performance

Under soft-label supervision, ALBERT achieved weighted F1=0.682 (95% CI: 0.660–0.705) and macro AUC=0.851 (95% CI: 0.836–0.864) in the Regular condition. Relative to hard-label baselines from Xu et al. ^16^, weighted F1 was lower by 0.163 in the Regular setting and 0.179 in the Augmented setting (Table 3).

**Table 3.** Soft-Label vs. Hard-Label Performance (3-Class, ALBERT)

| Supervision | Input | Overall |  | Per-Class Recall |  |  | Soft Metrics<sup>b</sup> |  |
| --- | --- | --- | --- | --- | --- | --- | --- | --- |
|  |  | wF1 | AUC | Neg | Neu | Pos | Soft CE | Brier |
| Hard<sup>a</sup> | Regular | 0.8457 | 0.8992 | — | — | — | — | — |
| Hard<sup>a</sup> | Augmented | 0.8598 | 0.9284 | 0.9142 | 0.6856 | 0.5545 | — | — |
| Soft | Regular | 0.6823 | 0.8505 | 0.9658 | 0.4583 | 0.3133 | 0.8408 | 0.0577 |
| Soft | Augmented | 0.6809 | 0.8526 | 0.9611 | 0.4686 | 0.2733 | 0.8401 | 0.0575 |
| $\Delta$ (Soft–Hard, reg.) | | −0.163 | −0.049 | +0.052 | −0.227 | — | — | — |
| $\Delta$ (Soft–Hard, aug.) | | −0.179 | −0.076 | +0.047 | −0.217 | −0.281 | — | — |

<sup>a</sup> Hard-label
results from Xu et al.<sup>16</sup> using the same dataset, encoder (albert-base-v2), and 60/20/20 split. <sup>b</sup> Soft CE = soft cross-entropy;<sup>28</sup> Brier score<sup>21</sup> evaluated against the full AI probability distribution. Both available only for soft-label conditions. Neg, Neu, Pos = per-class recall. Hard Regular per-class recall was not reported in Xu et al.<sup>16</sup>

Per-class recall showed a strong skew toward Negative predictions. In both soft-label conditions, Negative recall exceeded 0.96, whereas Neutral recall remained below 0.47 and Positive recall below 0.32. This pattern is consistent with the documented severity shift in the LLM annotator ^13^ and indicates that soft-label training did not recover human-aligned class boundaries. Token augmentation did not improve soft-label performance. Weighted F1 changed by −0.001 and macro AUC by +0.002 between the Regular and Augmented soft-label models.

#### 3.2.3 Soft evaluation metrics

When evaluated against the LLM’s own probability distributions rather than human labels, the soft-trained models achieved low Brier scores (Regular: 0.058, 95% CI: 0.054–0.062; Augmented: 0.057, 95% CI: 0.053– 0.062) and soft cross-entropy values of 0.841 (95% CI: 0.831–0.852) and 0.840 (95% CI: 0.830–0.851), respectively. Thus, the models reproduced the LLM probability landscape more closely than they matched human emotion labels.

In a supplementary 4-class soft-label analysis using the same encoder and training protocol, weighted F1 was 0.633 and Positive F1 was 0.076, indicating that 3-class collapse improved but did not eliminate the performance limitations of soft-label supervision in this setting.

## 4 DISCUSSION

Taken together, the 2 studies distinguish between 2 uses of LLM annotation infrastructure: as a source of scalable proxy burden labels for multi-task modeling, and as a source of distributional supervision for emotion classification. The results support the feasibility of modeling multidimensional burden-related signals from forum text, but they also show that auxiliary task design and label provenance strongly affect downstream performance.

### 4.1 Multi-Dimensional Burden Signal Modeling from Forum Text

The Composite model showed that a single ALBERT encoder could learn both a continuous LLM-derived burden score (*R*^2^=0.446; RMSE=13.47 on a 0–100 scale) and a binary high-need proxy flag (weighted F1=0.810; recall=0.94) from unstructured cancer peer-support text. Relative to prior work focused primarily on sentiment or emotion classification, ^29,30^ this extends automated monitoring toward multidimensional burden profiling. Because the burden targets were LLM-generated proxy annotations rather than validated clinical measures, these findings should be interpreted as evidence of learnability and internal coherence of the annotation scheme rather than validation of psychosocial burden measurement.

Performance differed across subscales. Cost burden showed the strongest classification performance (F1=0.852), whereas harm was weakest (F1=0.531). One plausible explanation is lexical distinctiveness: cost burden is often expressed with relatively specific financial terms, whereas harm overlaps more with broad negative affect in cancer support narratives. This gradient suggests differing levels of readiness across burden dimensions. Cost burden (F1=0.852) may be suitable for automated routing with periodic audit in research settings. Mid-tier subscales (support/coping, treatment burden, benefit; F1=0.62–0.69) are better suited to human-in-the-loop workflows where model outputs prioritize moderator attention. Low-tier subscales (harm, uncertainty; F1*<*0.59) should be treated as supplementary screening signals requiring human primary review.

From an informatics perspective, the results suggest that a relatively small encoder (12M parameters) can approximate multidimensional burden-related signals extracted by an LLM at lower inference cost. The composite score and high-need flag may therefore be useful for forum-level trend monitoring or moderator prioritization in research settings. However, prospective validation against established patient-reported outcome instruments, such as COST or FACT-G, remains necessary before clinical deployment or decision-support use.

### 4.2 Auxiliary Tasks Can Compete Rather Than Complement

Adding speaker role and cancer type as auxiliary prediction heads consistently reduced primary HEOR performance. The learned Kendall weights offer one optimization-based explanation: role prediction, an easier task (F1 *>* 0.91), received 35%–44% of the normalized task-weight budget, whereas cancer type received little weight and its own performance also declined as the number of heads increased. These findings suggest that, in this setting, easy auxiliary tasks can dominate shared optimization rather than improve the primary representation.

This pattern also clarifies the difference between using contextual variables as inputs versus as prediction targets. In Xu et al., ^16^ role and cancer-type information modestly improved emotion classification when added as input tokens. Here, the same information degraded burden modeling when implemented as auxiliary heads. Token augmentation enriches the input without creating additional supervised objectives, whereas auxiliary heads introduce optimization trade-offs within a shared encoder. When contextual metadata are available, input-side integration may therefore be preferable to output-side integration unless the auxiliary task is closely matched to the primary objective.

The preliminary failure of naive equal weighting in Composite+RC, which produced negative *R*^2^ values, further suggests that learned loss balancing is necessary when regression and classification tasks are optimized together, but not sufficient to eliminate competition among heterogeneous tasks.

### 4.3 Soft-Label Supervision Depends on Label Quality

Soft-label supervision reduced weighted F1 by 0.16 relative to hard-label baselines, with performance characterized by very high Negative recall (0.97) but poor Neutral and Positive recall. When evaluated against the LLM’s own probability distributions, the soft-trained models achieved low Brier scores, indicating that they reproduced the teacher distribution more closely than they aligned with human emotion labels. In other words, the soft-label objective preserved the annotator’s probability landscape, including its apparent severity shift.

This finding has practical implications for the growing use of LLM annotations in health NLP. ^10,31^ Soft labels are not inherently undesirable: when the teacher is well calibrated and the target of interest matches the teacher distribution, soft supervision can preserve useful uncertainty information. ^12,32^ In our setting, however, the LLM-derived distributions did not align well with human-labeled emotion categories, and soft-label training underperformed hard-label training accordingly. This result is consistent with broader work on label noise and teacher-student misalignment.^33,34^

The interaction with token augmentation is also informative. Xu et al. ^16^ found a modest gain from role and cancer-type token prepending under hard-label supervision, whereas the same augmentation had essentially no effect under soft-label training. This suggests that augmentation benefits may depend on label quality: when the supervision signal is biased or weakly aligned with the evaluation target, additional contextual features may not recover performance. A parallel finding emerged in a drug-review domain, where aspect-level LLM annotations required explicit fusion weighting to overcome ordinal label noise. ^35^ More generally, augmentation strategies should be evaluated jointly with annotation quality rather than as architecture choices in isolation.

### 4.4 Practical Implications

Several practical implications follow from these results. For burden modeling, the composite-only MTL configuration appears preferable to variants with auxiliary prediction heads. For emotion classification, hard-label supervision remained the safer choice in this setting, whereas LLM-derived probability distributions should be calibrated and audited before being used as training targets. More broadly, auxiliary tasks should be empirically justified rather than assumed to help, and LLM annotation pipelines should be evaluated not only for coverage and convenience but also for calibration, class balance, and agreement with the intended ground truth.

### 4.5 Limitations and Future Directions

This study has several limitations. First, all experiments used 1 English-language cancer peer-support dataset, ^17^ so generalizability to other platforms, conditions, or languages is unknown. Second, all AI labels were generated by a single LLM (GPT-4o-mini), ^19^ and alternative prompting strategies or multi-annotator LLM designs^36^ may yield different results. Third, the HEOR subscale labels lacked human validation and were defined by the LLM annotation rubric rather than established patient-reported outcome instruments such as COST or FACT-G. Accordingly, Study 1 should be interpreted as modeling an LLM-defined psychosocial burden ontology rather than validating a clinically established burden instrument. Fourth, we evaluated a single encoder architecture (ALBERT), and other model families may exhibit different task-interference patterns. Fifth, we did not test hybrid supervision strategies that combine hard human labels with soft AI distributions. Sixth, temporal stability and cross-platform transfer were not assessed.

Future work should evaluate dual-supervision approaches that anchor decision boundaries with human labels while incorporating calibrated soft distributions, prospectively validate burden dimensions against established patient-reported outcome measures, and compare alternative task-weighting strategies such as GradNorm and PCGrad to uncertainty weighting.

## 5 CONCLUSION

Multi-task learning supported the feasibility of modeling multidimensional psychosocial burden-related signals from cancer peer-support text, with the composite-only configuration achieving *R*^2^=0.446 for burden regression and weighted F1=0.810 for high-need screening. Adding auxiliary speaker-role and cancer-type prediction heads reduced primary-task performance, suggesting that auxiliary objectives can compete with rather than support the main burden tasks in a shared encoder. In parallel, LLM-derived soft-label supervision reduced emotion-classification performance by 0.16 weighted F1 relative to hard-label baselines, and token augmentation did not mitigate this gap.

These findings have 2 implications for health informatics. First, when contextual metadata are available, input-side integration may be preferable to auxiliary prediction heads for supporting primary text-classification tasks. Second, LLM-derived probability distributions should be calibrated and audited before use as training targets. Overall, composite-only MTL may support future human-in-the-loop monitoring workflows for patient-generated health data, but prospective validation of the burden constructs remains necessary before clinical deployment.

## Data Availability

The dataset analyzed in this study is publicly available. The Mental Health Insights: Vulnerable Cancer Survivors and Caregivers dataset is available in the Mendeley Data repository (doi:10.17632/69dcnv2gzd.1) under a CC BY 4.0 license and is mirrored on Kaggle and GitHub. LLM-derived annotations, train/validation/test split assignments, and analysis code will be made publicly available upon publication.

https://data.mendeley.com/datasets/69dcnv2gzd/1

## DATA AVAILABILITY STATEMENT

The dataset analyzed in this study is publicly available. The *Mental Health Insights: Vulnerable Cancer Survivors and Caregivers* dataset is available in the Mendeley Data repository (doi:10.17632/69dcnv2gzd.1) under a CC BY 4.0 license and is mirrored on Kaggle and GitHub. LLM-derived annotations, train/validation/test split assignments, and analysis code will be made publicly available upon publication.

## ACKNOWLEDGMENTS

The authors utilized the ALBERT pretrained language model via the Hugging Face Transformers library. LLM annotations were generated using the OpenAI API (GPT-4o-mini). During the preparation of this manuscript, the authors used Claude (Anthropic, San Francisco, CA, USA) for language editing and manuscript refinement. The authors reviewed and edited all AI-assisted output and take full responsibility for the content of the publication.

## FUNDING

This research received no external funding.

## COMPETING INTERESTS

The authors declare no competing interests.

## AUTHOR CONTRIBUTIONS (CREDIT)

Y.C. and Y.Z. contributed equally to this work. Z.W. conceived and supervised the study. Y.C. and X.S. developed and implemented the MTL framework. Y.Z. and Y.L. implemented the soft-label experiments. Z.D. contributed to data preprocessing and annotation pipeline development. Z.W. performed the statistical analysis and wrote the manuscript. All authors reviewed and approved the final manuscript.

## Notes

### Competing Interest Statement

The authors have declared no competing interest.

### Funding Statement

This study did not receive any funding

### Author Declarations

The study used (or will use) ONLY openly available human data that were originally located at: the Mendeley Data repository (doi:10.17632/69dcnv2gzd.1) under a CC BY 4.0 license and is mirrored on Kaggle and GitHub.

## REFERENCES

1. Wagle NS, Nogueira L, Devasia TP, Mariotto AB, Yabroff KR, Islami F, et al. Cancer treatment and survivorship statistics, 2025. CA: A Cancer Journal for Clinicians. 2025;75(4):308–40.

2. Lordon RJ, Mikles SP, Kneale L, Evans HL, Munson SA, Backonja U, et al. How patient-generated health data and patient-reported outcomes affect patient-clinician relationships: A systematic review. Health Informatics Journal. 2020.

3. Stetson PD, McCleary NJ, Osterman T, Ramchandran K, Tevaarwerk A, Wong T, et al. Adoption of Patient-Generated Health Data in Oncology: A Report From the NCCN EHR Oncology Advisory Group. Journal of the National Comprehensive Cancer Network. 2022;20(13).

4. Watanabe T, Yada S, Aramaki E, Yajima H, Kizaki H, Hori S. Extracting Multiple Worries From Breast Cancer Patient Blogs Using Multilabel Classification With BERT. JMIR Cancer. 2022;8(2):e37840.

5. Zhang Z, Liew K, Kuijer R, She WJ, Yada S, Wakamiya S, et al. Differing Content and Language Based on Poster-Patient Relationships on Weibo: Text Classification, Sentiment Analysis, and Topic Modeling of Posts on Breast Cancer. JMIR Cancer. 2024;10:e51332.

6. Edara DC, Vanukuri LP, Sistla V, Kolli VKK. Sentiment analysis and text categorization of cancer medical records with LSTM. Journal of Ambient Intelligence and Humanized Computing. 2023;14:5309–25.

7. American Association for Cancer Research. Supporting Cancer Patients and Survivors. American Association for Cancer Research; 2025. Available from: https://cancerprogressreport.aacr.org/progress/cpr25-contents/cpr25-supporting-cancer-patients-and-survivors/.

8. Caruana R. Multitask Learning. Machine Learning. 1997;28:41–75.

9. Kendall A, Gal Y, Cipolla R. Multi-task Learning Using Uncertainty to Weigh Losses for Scene Geometry and Semantics. In: 2018 IEEE/CVF Conference on Computer Vision and Pattern Recognition; 2018. p. 7482–91.

10. Gilardi F, Alizadeh M, Kubli M. ChatGPT Outperforms Crowd-Workers for Text-Annotation Tasks. Proceedings of the National Academy of Sciences of the United States of America. 2023;120(30):e2305016120.

11. Yao T, Foo E, Binnewies S. CALLM: Context-Aware Emotion Analysis in Cancer Survivors Using LLMs and Retrieval-Augmented Mobile Diaries; 2025. Available from: https://arxiv.org/abs/2503.10707.

12. Uma A, Fornaciari T, Muresan S, Plank B, Poesio M. Learning from disagreement: A survey. Journal of Artificial Intelligence Research. 2021;72:1385–418.

13. Lu J, Ma K, Wang K, Xiao K, Lee RKW, Xu B, et al. Is LLM an Overconfident Judge? Unveiling the Capabilities of LLMs in Detecting Offensive Language with Annotation Disagreement. In: Findings of the Association for Computational Linguistics: ACL 2025. Association for Computational Linguistics; 2025. p. 5609–26. Available from: https://aclanthology.org/2025.findings-acl.293/.

14. Koo R, Lee M, Raheja V, Park JI, Kim ZM, Kang D. Benchmarking Cognitive Biases in Large Language Models as Evaluators. In: Findings of the Association for Computational Linguistics: ACL 2024. Association for Computational Linguistics; 2024. p. 517–45. Available from: https://aclanthology.org/2024.findings-acl.29/.

15. Wang Z, Cao Y, Xu S, Ding Z, Hao Y, Mao Qinkai T Qi, et al. Digital Phenotyping of Psychosocial Distress in Cancer Support Forums: A Statistically Rigorous Benchmark of Ordinal Sentiment Models; 2026. Under Review.

16. Xu S, Wang Z, Wang H, Ding Z, Zou Y, Cao Y. LLM-Based Annotation and Token-Augmented Modeling for Emotional Tone Classification in Online Cancer Peer-Support Posts; 2026. Under review.

17. Orchi IH, Tabassum N, Hossain J, Tajrin S, Alam I. Mental Health Insights: Vulnerable Cancer Survivors & Caregivers Data. Mendeley Data; 2023. Available from: https://data.mendeley.com/datasets/69dcnv2gzd/1.

18. Lan Z, Chen M, Goodman S, Gimpel K, Sharma P, Soricut R. ALBERT: A Lite BERT for Self-Supervised Learning of Language Representations. In: Proceedings of the 8th International Conference on Learning Representations (ICLR); 2020. Available from: https://openreview.net/forum?id=H1eA7AEtvS.

19. OpenAI. OpenAI API (gpt-4o-mini); 2024. Computer software. https://platform.openai.com.

20. Lin J. Divergence Measures Based on the Shannon Entropy. IEEE Transactions on Information Theory. 1991;37(1):145–51.

21. Brier GW. Verification of Forecasts Expressed in Terms of Probability. Monthly Weather Review. 1950;78(1):1–3.

22. Sokolova M, Lapalme G. A systematic analysis of performance measures for classification tasks. Information Processing & Management. 2009;45(4):427–37.

23. Gneiting T, Katzfuss M. Probabilistic Forecasting. Annual Review of Statistics and Its Application. 2014;1:125–51.

24. Efron B, Tibshirani RJ. An Introduction to the Bootstrap. New York: Chapman & Hall/CRC; 1994.

25. Paszke A, Gross S, Massa F, Lerer A, Bradbury J, Chanan G, et al. PyTorch: An Imperative Style, High-Performance Deep Learning Library. In: Advances in Neural Information Processing Systems 32; 2019. p. 8026–37.

26. Wolf T, Debut L, Sanh V, Chaumond J, Delangue C, Moi A, et al. Transformers: State-of-the-Art Natural Language Processing. In: Proceedings of the 2020 Conference on Empirical Methods in Natural Language Processing: System Demonstrations. Association for Computational Linguistics; 2020. p. 38–45. Available from: https://aclanthology.org/2020.emnlp-demos.6/.

27. Shannon CE. A Mathematical Theory of Communication. Bell System Technical Journal. 1948;27(3):379–423.

28. Szegedy C, Vanhoucke V, Ioffe S, Shlens J, Wojna Z. Re-thinking the Inception Architecture for Computer Vision. In: Proceedings of the IEEE Conference on Computer Vision and Pattern Recognition (CVPR); 2016. p. 2818–26.

29. Calvo RA, Milne DN, Hussain MS, Christensen H. Natural language processing in mental health applications using non-clinical texts. Natural Language Engineering. 2017;23(5):649–85.

30. Shatte ABR, Hutchinson DM, Teague SJ. Machine learning in mental health: A scoping review of methods and applications. Psychological Medicine. 2019;49(9):1426–48.

31. Mellon J, Bailey J, Scott R, Breckwoldt J, Miori M, Schmedeman P. Do AIs Know What the Most Important Issue Is? Using Language Models to Code Open-text Social Survey Responses at Scale. Research & Politics. 2024;11(1):1–10.

32. Niculescu-Mizil A, Caruana R. Predicting Good Probabilities with Supervised Learning. In: Proceedings of the 22nd International Conference on Machine Learning. ACM; 2005. p. 625–32.

33. Frénay B, Verleysen M. Classification in the Presence of Label Noise: A Survey. IEEE Transactions on Neural Networks and Learning Systems. 2014;25(5):845–69.

34. Northcutt C, Jiang L, Chuang I. Confident Learning: Estimating Uncertainty in Dataset Labels. Journal of Artificial Intelligence Research. 2021;70:1373–411.

35. Liu J. Learning to Fuse Aspect-Level LLM Annotations for Low-Quality Ordinal Sentiment Supervision; 2026. SSRN Working Paper, posted December 24, 2025. Available at SSRN.

36. Argyle LP, Busby EC, Fulda N, Gubler JR, Rytting C, Wingate D. Out of One, Many: Using Language Models to Simulate Human Samples. Political Analysis. 2023;31(3):337–51.

